# The Evolving Landscape of Substance Use Disorder Mortality in the United States: A Spatiotemporal Analysis of Emerging Hotspots and Vulnerable Populations (2005-2020)

**DOI:** 10.1101/2024.07.08.24310099

**Authors:** Santiago Escobar, Neil J. MacKinon, Preshit Ambade, Zach Hoffman, Diego F Cuadros

**Author notes:** Corresponding author: Diego Cuadros.

## Abstract

The escalating Substance Use Disorder (SUD) crisis in the U.S., marked by a significant rise in mortality since 1999, underscores the urgent need for a comprehensive analysis of its spatiotemporal dynamics. This study aims to elucidate the heterogeneous geospatial distribution of SUD mortality, identifying specific locations where vulnerable communities face heightened risk. By examining SUD mortality data from the CDC for the period 2005-2020, we applied scan statistics to delineate temporal and geospatial clusters of elevated SUD-related deaths, further dissecting these patterns across racial subpopulations and regions. Our findings reveal 27 distinct clusters nation-wide, predominantly emerging post-2013 and persisting until 2020, indicating a shifting epicenter of the epidemic. Notably, the white subpopulation was associated with 26 clusters, closely mirroring the broader national trends, yet with a pronounced concentration in the eastern U.S. Conversely, the black subpopulation demonstrated a different pattern, with 17 clusters arising between 2013 and 2020, primarily post-2015, suggesting a temporal and spatial divergence in the impact of the epidemic across racial subpopulations. This analysis not only highlights the critical need for targeted public health interventions and policies but also calls for continued surveillance to monitor and mitigate the evolving SUD crisis. By understanding the complex spatiotemporal and racial variations in SUD mortality, we can better allocate resources, develop effective prevention strategies, and support the communities most in need.

## 1 Introduction

Substance use disorder (SUD) is defined as a treatable mental disorder that affects a person’s ability to control their use of substances, understood as compounds that can potentially be abused for recreational purposes due to their psychoactive nature.[1] This condition has been rising for over three decades, claiming over 900,000 lives in the United States (US) since 1999 and constituting a national epidemic.[2] Deaths from SUD have increased more than any other cause of death in this period, with SUD-related deaths surpassing motor vehicle traffic as the most frequent unintentional injury-related death cause in 2012 and staying on top ever since.[3] The spatial distribution of the SUD epidemic has shown a heterogeneous pattern, with some areas exhibiting increased risk. The West and Midwest regions of the US have shown clusters of SUD mortality,[4] with temporal and social dynamics showing regional differences accounting for the local variation of the national epidemic. Temporarily, there are four distinct but overlapping waves of the epidemic.[5] The first wave, characterized by prescription opioids, occurred between 2000 and 2016. A rise in heroin-related deaths marked the second wave, beginning in 2007 and surpassing prescription-related deaths in 2015. The third wave is linked to synthetic opioids, which exhibited a steady increase from 2013 to 2018. The fourth wave is associated with a rise in polydrug use following the COVID-19 pandemic and could be considered to still be ongoing.

The racial differences are prominent in SUDs, with higher overall mortality among whites but with a sharp rise among black males post-2014.[6] In addition, the urban-rural divide has also created geographical differences: rural areas present a higher percentage of the white population, higher unemployment rates, and a greater percentage of specialized opioid prescribers, such as surgeons and oncologists,[7] while lacking resources to treat SUD.[8] These differences observed in the SUD epidemic can be attributed to supply and demand: the availability of certain substances over limited geographical areas may have caused variations in the observed mortality trends over time.[9]

Previous studies,[4] conducted a spatial analysis of the data up to 2017, highlighting geospatial hotspots emphasizing vulnerable populations. Extending this analysis further, this research seeks to explore the temporal dynamics of the spatial structure of the SUD crisis by examining the spatiotemporal dynamics of SUD mortality rates, including data until 2020.

Against this background, the primary aim of this study is to unravel the intricate spatiotemporal dynamics of SUD in the US over a period from 2005 to 2020. This study aims to delineate the evolving patterns of SUD mortality across different racial groups and geographical landscapes, offering insights into the local micro-epidemics that collectively shape the national crisis. By employing spatiotemporal clustering analysis, our study seeks to identify and characterize specific areas and populations most afflicted by SUD-related mortality, providing a detailed map of the epidemic’s progression over time. This study will pave the way for understanding the complex temporal and spatial interplay among socioeconomic, demographic, and spatial factors driving the SUD epidemic, with the goal of informing more effective prevention, treatment, and policy strategies.

## 2 Methods

### 2.1 Study design

A longitudinal ecological study was conducted to identify the spatiotemporal dynamics of SUD mortality across the contiguous U.S from 2005 to 2020. By employing retrospective spatiotemporal scan statistics, this study sought to identify the spatial and temporal structure followed by the SUD epidemic, analyzing the behavior of SUD – related deaths across the general population, as well as across the White, Black, Rural, and Urban subpopulations.

### 2.2 Data sources

SUD mortality data was collected from the US Vital Statistics System restricted-use micro-data mortality files for the period of January 2005 to December 2020,[10] the filtered data included date and county of death, decedent’s demographic characteristics (sex and race) and the International Classification of Diseases, 10th Revision (ICD-10) code for the cause of death.[11] Individuals aged 5 to 84 years were included, and drug overdoses were identified as those with ICD-10 codes indicating unintentional substance poisoning (cause of death codes: X40, X41, X42, X43, X44) to estimate SUD mortality rates. These ICD-10 codes included deaths caused by the following substances: heroin, methadone, cocaine, other opioids, synthetic narcotics, and unspecified narcotics.

Population at risk was estimated by using the latest county-level population estimates from the U.S Census Population and Housing Unit Estimates Tables for each of the years of our study period.[12] Data was filtered by subpopulation, obtaining yearly population estimates by demographic characteristics (sex and race). Further, counties were classified as urban or rural following the National Center for Health Statistics (NCHS) 2013 Urban-Rural classification schemes [13]: Large central metro, Large fringe metro, Medium metro, and Small metro counties were considered urban, while Micropolitan and Noncore counties were considered rural. To delineate the heterogeneity in the estimates, county-level mortality rates are reported for the general population, encompassing the three racial categories reported in the mortality files of White, Black, and Others. Further, separate estimates for White, Black, Urban, and Rural subpopulations are also reported.

### 2.3 Spatiotemporal Clustering analysis

A county-level spatiotemporal clustering analysis was conducted to identify geographical clusters of high numbers of SUD-related deaths that persist for at least two or more years using Kulldorff’s spatial scan statistics [14] implemented in the SaTScan software. The analysis was performed for the general population, as well as for the White and Black subpopulations. Scan statistics are widely used for cluster detection in epidemiology [15-18], social sciences,[19] crime mapping,[20] and, very recently, in mental health,[21] among other applications. A detailed description of the spatial scan statistics is provided elsewhere.[14, 17] Briefly, Kulldorff’s scan statistics are used to detect high-risk spatiotemporal clusters of cases (i.e. SUD-related deaths) by gradually scanning a cylindrical window, with a base corresponding to space and the height corresponding to time. Cylinders of varying radii spanned the study region and time-period to identify areas where SUD-related deaths were clustered. The cylinder varied continuously in both location and radius, thus creating and testing a very large number of distinct potential clusters. A log-likelihood ratio of the test scan statistic is constructed based on the actual number of occurrences and the expected number of occurrences inside and outside the cylinder. Clusters with a P < 0.05, calculated through Monte Carlo simulations (using the default value of 999 iterations), were identified as statistically significant clusters of SUD-related deaths. After a cluster was identified, the strength of the clustering was estimated using the relative risk (RR) of cases within the cluster versus outside the cluster. The number of SUD-related deaths from the complete dataset (2005-2020) was analyzed at the county level using Kulldorff’s spatial scan statistics with a Poisson model with the overall population size by county included as an offset. SUD mortality rates are reported as the number of deaths per 10,000 people.

### 2.4 Analyzing the Urban and Rural divide in the epidemic

To assess the distribution of the epidemic across urban and rural areas, bivariate choropleth maps were produced for the urban/rural status of a county and if it was classified as a cluster for the general population at some point in the study period. To illustrate the statistical relationship established between these variables and their differences in the East and West of the country, proportionate bar graphs were produced using the R package ggplot2.[22] Lastly, contingency tables were created for the urban/rural status of a county and whether it was classified as a cluster for the general population at some point in the study period, these tables were used to estimate the Odds Ratio (OR) for a rural county being classified as a cluster compared to a urban county. ORs were estimated for the whole country, as well as counties to the East and to the West of the country.

### 2.5 Temporal dynamics across subpopulations

Mean mortality values were estimated across the country by year for the general population, as well as for the White, Black, Urban, and Rural subpopulations. The mean mortality estimates were used to produce temporal line graphs representing the variability in mortality trends across time by subpopulation.

## 3 Results

### 3.1 General results

Deaths by unintentional drug overdose in the contiguous U.S. from 2005 to 2020 resulted in 665,341 cases. The estimated average SUD mortality rate during this period in the total population was 1.13 per 10,000 people per year. The state with the highest mortality rate during this period was West Virginia, with a mortality rate of 2.37 per 10,000 people per year, followed by Kentucky (2.21 per 10,000), Ohio (2.01 per 10,000), Rhode Island (1.85 per 10,000), and Tennessee (1.75 per 10,000). The estimated SUD mortality rate in the White subpopulation was 1.22 per 10,000 people per year, whereas the mortality rate in the Black subpopulation was 1.11.

### 3.2 Spatiotemporal clustering analysis for the general population

Spatiotemporal SaTScan analysis identified 27 significant spatiotemporal clusters that started emerging in 2005 (Figure 1). The estimated mortality rate within the clusters was 1.51 per 10,000 people per year compared with a mortality rate of 0.86 per 10,000 people per year estimated in the areas outside of the identified clusters. One cluster emerged in the state of Washington in 2005, lasting until 2009 (average mortality rate 1.52 per 10.000), a second early cluster emerged in the state of Oklahoma in 2009, lasting until 2016 (average mortality rate 1.80 per 10.000). The 25 remaining clusters are divided into two main temporal groups: ten clusters starting from 2013 to 2015 and lasting until the end of the study period (average mortality rate 2.00 per 10.000), and 15 clusters starting from 2016 to 2018 and lasting until the end of the study period (average mortality rate 1.15 per 10.000). Broadly, most of the clusters emerging before 2016 were located in the western part of the country, whereas most of the clusters emerging after 2016 were located in the eastern part of the country.

**Figure 1:**
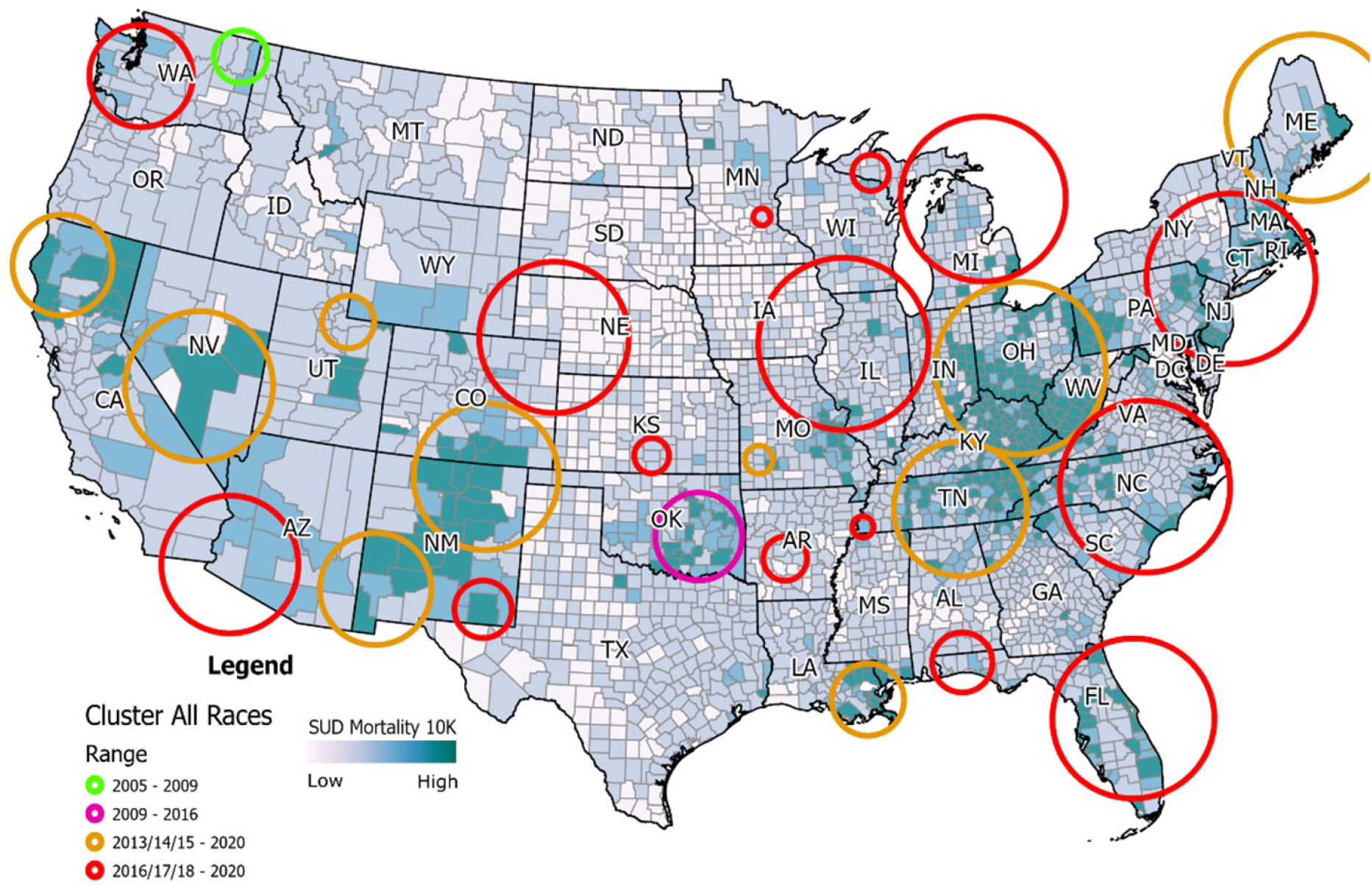
Spatiotemporal Clustering of SUD-Related Deaths in the Contiguous United States (2005-2020). This map shows significant spatiotemporal clusters of SUD-related deaths in the contiguous United States from 2005 to 2020, identified using Kulldorff’s spatial scan statistics. Early clusters appeared in Washington (2005-2009) and Oklahoma (2009-2016). From 2013 onward, clusters emerged in the western US, shifting to the eastern US post-2016. The mortality rate within clusters was 1.51 per 10,000 people per year, compared to 0.86 per 10,000 in non-cluster areas, highlighting regional variations and the shifting epicenter of the SUD epidemic.

### 3.3 Spatiotemporal clustering analysis for the subpopulations

Spatiotemporal clusters of high SUD mortality were also detected for the White and Black subpopulations, using the same analysis and parameters. For the White subpopulation, 26 significant clusters were reported, with a spatial distribution that exhibits similar structure as observed for the general population (Figure 2A). The estimated mortality rate within the clusters was 1.83 per 10,000 people per year compared with a mortality rate of 0.90 per 10,000 people per year estimated in the areas outside of the identified clusters. One cluster emerged in the state of Washington between the years 2005 to 2009 (average mortality rate 1.62 per 10.000), two more early clusters showed up in the states of California and Oklahoma from 2009 to 2016 (average mortality rate 1.97 per 10.000). Eleven clusters starting from 2013 to 2015 and lasting until the end of the study period were reported (average mortality rate 2.12 per 10.000), and 12 clusters starting from 2016 to 2018 and lasting until the end of the study period (average mortality rate 1.53 per 10.000). Similar to the results reported for the general population, most of the clusters pre-2016 emerged in the western part of the country, whereas the clusters post-2016 emerged in the eastern part of the country.

**Figure 2:**
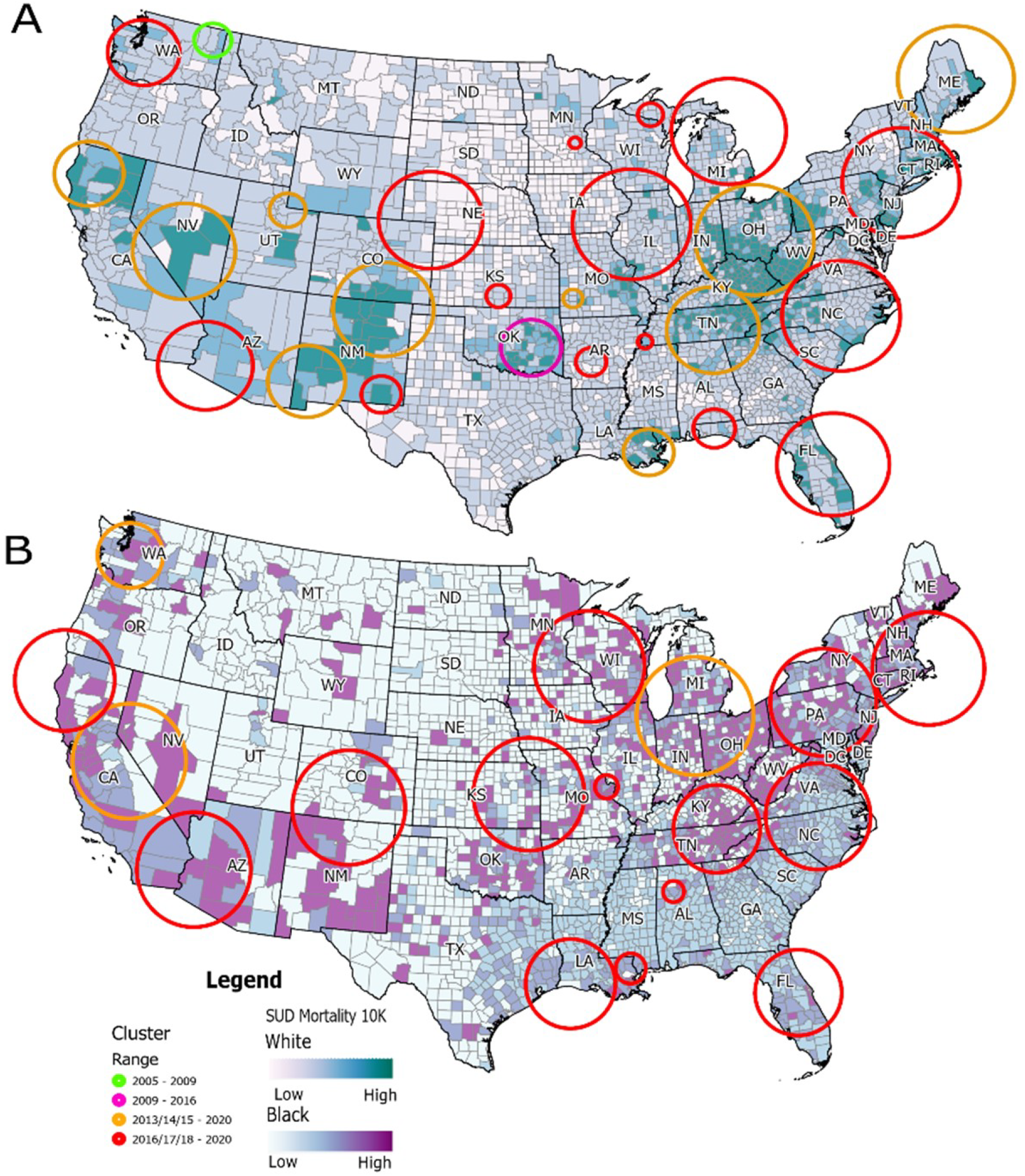
Spatiotemporal Clusters of SUD Mortality by Race in the US (2005-2020). This figure displays spatiotemporal clusters of SUD-related deaths by race in the contiguous United States from 2005 to 2020. A) Clusters for the White subpopulation. A total of 26 significant clusters were identified. Early clusters appeared in Washington (2005-2009) and Oklahoma (2009-2016), with additional clusters emerging in California. Post-2013, clusters proliferated in the western US, with a shift to the eastern US post-2016. The mortality rate within clusters for whites was 1.83 per 10,000 people per year, compared to 0.90 per 10,000 in non-cluster areas. B) Clusters for the Black subpopulation. Seventeen significant clusters were identified, all emerging between 2013 and 2018 and continuing until 2020. These clusters were predominantly located in the eastern US. The mortality rate within clusters for blacks was 1.66 per 10,000 people per year, compared to 0.81 per 10,000 in non-cluster areas. This reflects a later but rapid emergence of clusters in the Black subpopulation compared to the White subpopulation.

For the Black subpopulation, 17 significant clusters were identified, all of which started between 2013 and 2018 and continued to 2020 (Figure 2B). The mortality rate estimated inside of the counties identified as clusters was 1.66 per 10,000 people per year, while in the counties outside of clusters the estimated mortality rate was 0.81 per 10,000 people per year. Only three clusters were reported between 2013 and 2015 (average mortality rate 2.77 per 10.000), while the remaining 14 clusters were reported as emerging after 2016 (average mortality rate 1.28 per 10.000). These SUD clusters for Black subpopulation were predominantly located in the eastern part of the country.

### 3.4 SUD mortality in Urban and Rural areas

The estimated average SUD mortality rate for the study period was 1.30 per 10,000 people per year in the urban areas, compared to 1.03 per 10,000 people per year in the rural areas. The average mortality rate in urban counties classified as clusters was 1.61 per 10,000 people per year, while in urban counties not classified as clusters it was 0.97 per 10,000 people per year. For rural counties classified as clusters the average mortality rate was 1.43 per 10,000 people per year, while rural counties not classified as clusters had an average mortality rate of 0.81 per 10,000 people per year. It is important to note that due to the nature of the analysis, clusters can contain both urban and rural counties, but as it can be observed in Figure 3, clusters located to the West of the country are predominantly rural, whereas to the East of the country the proportion of urban and rural counties inside of clusters is relatively even.

**Figure 3:**
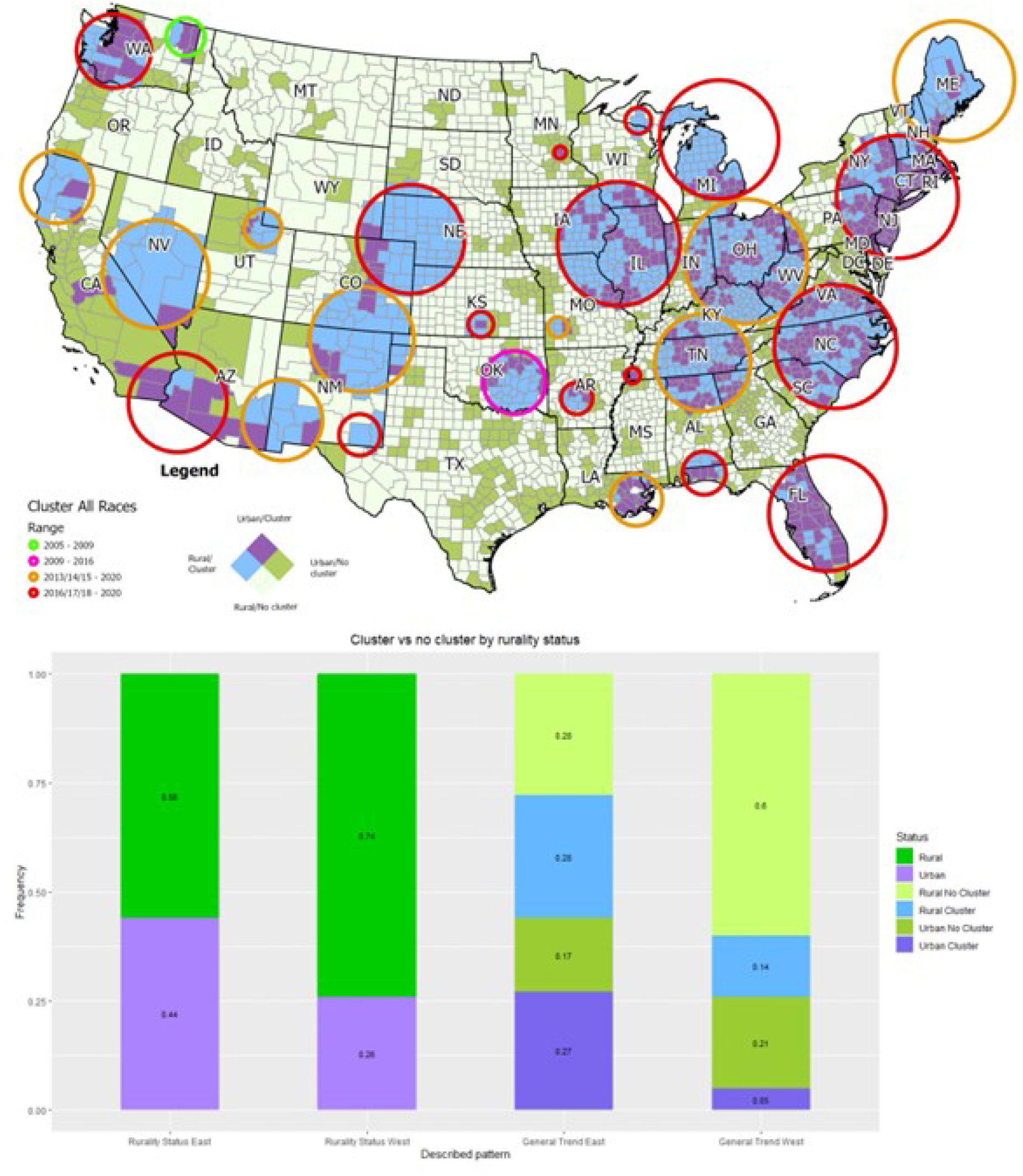
Urban-Rural Differences in SUD Mortality Clusters in the US (2005-2020). This figure illustrates the urban-rural differences in SUD-related mortality clusters in the contiguous United States from 2005 to 2020. A) Bivariate choropleth map showing urban and rural counties classified as clusters. Urban counties are predominantly found in clusters in the eastern US, whereas clusters in the western US are mostly rural. The map highlights the spatial distribution of high-risk areas, with urban areas showing higher cluster prevalence in the East and rural areas in the West. B) Bar chart depicting the proportion of urban and rural counties within identified clusters across the US. The chart demonstrates a higher likelihood of rural counties being classified as clusters in the western US, while urban counties are more prevalent in clusters in the eastern U.S.

The analysis of the data for the country in general revealed an OR of 0.57 for belonging to a cluster in rural counties compared to urban counties: Rural counties were 0.57 times as likely to be classified as clusters compared to urban counties, which implies that the presence of urban counties within clusters was almost twice as compared to rural counties. In terms of the urban-rural concentration of counties within a cluster, the East and West regions of the country showed a contrasting picture. Rural counties from the East were less likely to be in the SUD cluster compared to their urban counterparts (OR: 0.63), whereas in the West their likelihood was higher (OR:1.04).

### 3.5 Temporal dynamics by subpopulation

Cumulative mortality graphs were produced for each subpopulation, contrasting the mortality for the subpopulation, for the counties within the SUD clusters in that subpopulation, and for the counties outside of these clusters in that subpopulation (Figure 4). For the general population, there is a clear difference in the mortality observed inside of the clusters vs outside of the clusters, this contrast remains stable over time besides a sudden drop observed in 2008 for the mortality values inside of the clusters. Then, the estimated mortality observed outside of clusters drop below the estimated average mortality for the general population from 2012 onward. The temporal trend of the SUD-related mortality in the White subpopulation closely resembles the patterns observed for the general population, with a reduction in the estimated SUD-related mortality observed in 2008 within the identified clusters. In contrast, the SUD-mortality in the Black subpopulation showed a different temporal pattern: a homogeneous spatial structure resulted in the absence of clusters until 2013, when we observe the first clusters rapidly emerge across the country. The emergence of these clusters is associated to an increase in SUD-related deaths in the Black subpopulation when compared with the national averages. Likewise, from 2016 onwards, the mortality rate observed in Black surpasses that of the White subpopulation. It became the highest SUD mortality rate among the observed subpopulations, significantly spiking in 2018 and onwards.

**Figure 4:**
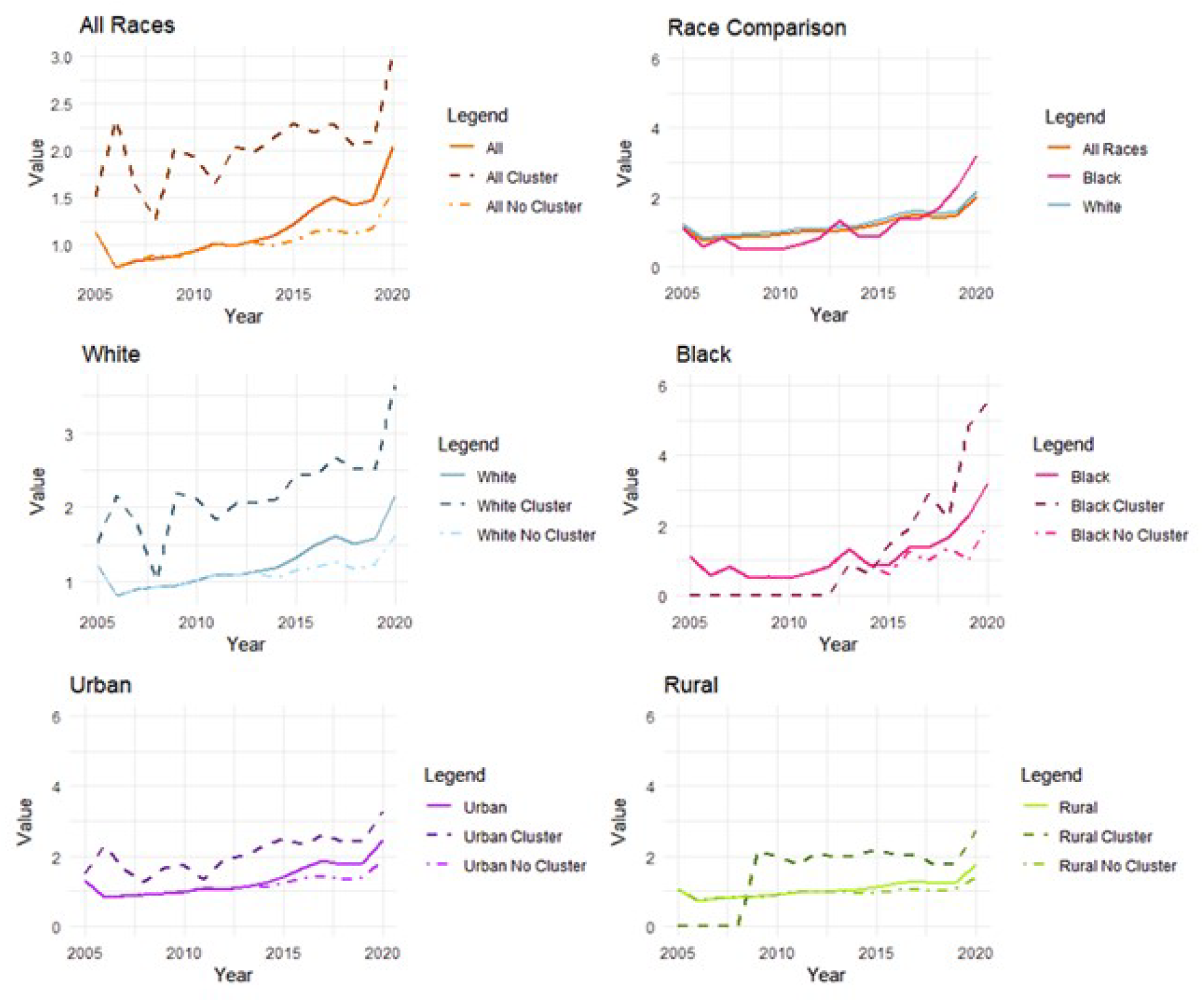
Temporal Trends in SUD Mortality by Subpopulation and Clusters (2005-2020). This figure presents the temporal trends in SUD-related mortality across different subpopulations in the contiguous United States from 2005 to 2020. Top panels, cumulative mortality graph for the general population (left), and comparative cumulative mortality across subpopulations (right). Middle panels, temporal trends for the White subpopulation (right), and temporal trends for the Black subpopulation (right). Bottom panels, temporal trends for the Urban subpopulation (left), and temporal trends for the rural population (right). These temporal graphs illustrate the evolving dynamics of the SUD epidemic, highlighting distinct temporal patterns and the emergence of clusters within different subpopulations over time.

A distinct geographical disparity between urban and rural areas was observed in the mortality rates within and outside the identified clusters, exhibiting temporal fluctuations. Yet, urban counties predominantly characterized the clusters over the study period. Urban clusters were identified early in the study timeline, displaying variable mortality rates in relation to urban national averages. In contrast, clusters within rural areas emerged post-2009, consistently demonstrating a stable divergence in mortality rates when juxtaposed against the rural national average.

## 4 Discussion

The spatiotemporal dynamics of the SUD epidemic in the US reveal the emergence of distinct localized micro-epidemics with unique characteristics. Using spatial scan statistics, clusters of elevated SUD mortality rates that exceeded expected values were identified. Initial clusters were detected in Washington (2005-2009) and Oklahoma (2009-2016). From 2013 to 2016, clusters proliferated in the western US, persisting until 2020, while clusters in the eastern US emerged from 2016 to 2018 and continued through the study period. Within these clusters, the mortality rate was 1.51 per 10,000 individuals annually, compared to 0.86 per 10,000 in non-cluster regions, highlighting the need for region-specific public health interventions.

Socioeconomic patterns driving the emergence of these localized micro-epidemics are still under investigation. Studies suggest that different substances targeting specific demographics have initiated distinct epidemic phases.[4, 9, 23] The initial phase involved prescription opioid misuse, followed by a surge in heroin-related deaths from 2007. The subsequent phase, characterized by fatalities from fentanyl and other synthetic opioids, marks a transition driven by heroin’s increased availability and the challenges of obtaining prescription opioids. Recently, synthetic opioids misrepresented as other drugs have caused a spike in fatalities due to their potency heroin distribution in the US shows regional distinctions, with Colombian-sourced heroin prevalent in the East and Mexican “black tar” heroin in the West.[24] Powdered heroin, common in the Northeast and Midwest, is more often adulterated with fentanyl than the solid “black tar” heroin in the West.[25] These patterns illustrate the complex progression of the SUD epidemic, driven by varying supply and demand across different regions.

Our analysis revealed distinct SUD mortality patterns among racial subpopulations. Spatiotemporal scan statistics showed that clusters in the white subpopulation appeared early, similar to the general population pattern, while clusters in the black subpopulation emerged later, starting in 2013 and persisting through the study period. Within these clusters, the white subpopulation had a higher mortality rate (1.83 per 10,000) compared to non-cluster areas (0.90 per 10,000). For the black subpopulation, the rate within clusters was 1.66 per 10,000, versus 0.81 per 10,000 outside. Notably, from 2016 onwards, the mortality rate among blacks surpassed that of whites, with a significant spike from 2018 onwards.

The early predominance of white mortality rates is linked to prescription opioid misuse, influenced by disparities in pain management access and systemic healthcare biases affecting black patients.[9, 26, 27] The later emergence of clusters among blacks aligns with the rise of illicit synthetic opioids, particularly fentanyl, as shown by studies in St. Louis, Missouri, and Massachusetts, highlighting racial inequities in opioid overdose deaths disproportionately affecting the black population.[28, 29] These patterns emphasize the need for further research into the interplay between increased mortality, social determinants of health, and substance availability impacts on these demographic groups.

The estimated average SUD mortality rate during the study period was 1.30 per 10,000 individuals annually in urban areas, compared to 1.03 per 10,000 in rural areas. A segmented analysis between the eastern and western regions reveals a stark contrast: rural counties in the East had a lower likelihood of being classified as clusters compared to urban counties, while in the West, rural counties were more likely to form clusters than their urban counterparts. This analysis highlights a predominantly rural composition of clusters in the West.

This geographic disparity, with the West experiencing a predominantly rural SUD epidemic, can be attributed to several factors. Early cluster formations in the West, accelerated opioid overdose death rates in rural versus urban counties prior to 2010,[30] and higher prescription opioid rates in these rural areas contribute to this trend.[31, 32] Rural-urban health disparities, compounded by the demographic and socioeconomic makeup of rural populations, characterized by a higher proportion of older adults with chronic pain and a significant non-Hispanic white demographic with greater access to opioid prescriptions,[33, 34] further explain the observed spatial distribution of SUD mortality. These findings underscore the complex interplay between geographic location, demographic characteristics, and access to healthcare resources in the unfolding of the SUD epidemic across the US.

Our research highlights that the national SUD epidemic is shaped by numerous localized micro-epidemics, necessitating targeted local interventions to mitigate the overall national burden. Precise and timely detection using fine spatial and temporal resolution is essential to identify populations at higher risk. These risks involve both demand-side factors, such as socioeconomic status, educational attainment, and employment rates, and supply-side dynamics, including the availability of specific substances. Demand-side factors are generally stable and spatially specific, while supply-side elements are more volatile, often shifting rapidly over time.[9] The introduction of unregulated substances into the illicit market significantly impacts SUD mortality rates. The infiltration of fentanyl adulterated or substituted heroin (FASH) into the US market has markedly increased mortality, underscoring the critical role of supply dynamics in the epidemic’s evolution.[35] The use of spatiotemporal surveillance systems supports the creation of Early Warning Systems (EWS) for populations at increased risk due to demographic characteristics and exposure to new, unregulated substances. Such systems have been implemented at regional, national, and international levels, with entities like the United Nations Office of Drug and Crime (UNDOC), the Inter-American Drug Abuse Control Commission, and the European Monitoring Centre for Drugs and Drug Addiction providing foundational guidelines for EWS development and implementation.[36]

The National Drug Early Warning System (NDEWS) in the US specializes in disseminating hotspot alerts that identify deviations from expected drug-related incident rates within defined temporal windows. NDEWS, along with the State and National Overdose Web (SNOW) and the Florida Drug-Related Outcomes Surveillance and Tracking System (FROST), exemplifies the US’s advanced surveillance efforts to monitor and respond to emerging drug threats.[37-39] Additional early warning programs include DOSE,[40] DAWN,[41] and SUDOR.[42] Despite these advancements, the spatial granularity of reported data often remains suboptimal for detailed risk assessments. Integrating broader datasets, including those from emergency departments, medical examiners, and hospital records, into existing surveillance frameworks can enhance the precision and utility of EWS for effectively addressing the SUD epidemic.

According to the CDC’s Guiding Principles, addressing the SUD crisis requires innovative strategies to prevent overdoses and related harms.[43] The SUD epidemic’s dynamics are influenced by both demand, driven by socioeconomic factors within communities, and supply, dictated by the availability of controlled substances. Effective interventions must address these socioeconomic determinants and implement strict substance control policies. Additionally, recognizing individual decision-making in substance consumption suggests the need for comprehensive support, including educational initiatives, Naloxone distribution, and the development of surveillance and EWS.

EWS have proven beneficial for both policymakers and individuals directly affected by SUD. In Canada, provinces like British Columbia [44] and Saskatchewan [45] have implemented drug alert systems that provide real-time updates on hazardous substances within localities. These alerts offer detailed information about drugs, including their appearance and recent overdose locations, aiding informed decision-making among substance users. British Columbia also offers Drug Checking services, enabling individuals to test substances for dangerous adulterants like fentanyl and benzodiazepines. These services enhance community safety by providing accurate drug supply information. Although potentially contentious, such approaches emphasize the importance of engaging with communities affected by SUD to mitigate new risks. By analyzing drug testing information, a clearer understanding of the controlled substances market is gained, contrasting significantly with insights from clinical records. This facilitates targeted interventions at both individual and policy levels, illustrating a holistic approach to addressing the complex challenges of the SUD epidemic.

The identification of spatiotemporal clusters and the differential impact of the SUD epidemic on urban versus rural areas provides critical insights for designing and implementing effective EWS. Understanding the spatiotemporal dynamics and identifying high-risk regions and populations allows EWS to deliver targeted alerts and information, enabling rapid, focused responses. For example, recognizing that rural areas in the West are more likely to form clusters can guide regional strategies such as deploying mobile Naloxone units or establishing localized drug checking services. Additionally, temporal analysis offers a framework for predicting potential future outbreaks, facilitating preemptive actions in emerging risk communities. A detailed understanding of the dynamics of the epidemic supports innovative SUD management and prevention approaches. Policymakers can use our findings on demand and supply characteristics to craft multifaceted strategies addressing both immediate substance availability risks and deeper socioeconomic drivers of substance use. This might involve educational campaigns tailored to specific community needs and contexts, along with policies improving healthcare access and support services. Applying our results to EWS design and other interventions promises a more proactive, informed public health approach, mitigating the impact of the SUD epidemic.

At the local level, our findings highlight the importance of community-specific interventions. For instance, the identification of higher cluster formation in rural areas of the West suggests the need for enhanced resources and services in these regions. Local health departments should consider deploying mobile units for Naloxone distribution and establishing localized drug checking services to reduce overdose fatalities. Additionally, targeted educational campaigns that address the specific needs and contexts of rural populations can raise awareness about the risks of synthetic opioids like fentanyl.

On a national scale, our study supports the integration of advanced surveillance systems, such as FROST, SNOW, DOSE, SUDOR, and DAWN, to provide timely and precise alerts about specific substance threats. These Early Warning Systems can facilitate the rapid deployment of resources to areas identified as high-risk, allowing for a more proactive response to emerging trends in substance use and overdoses. Likewise, our findings on racial disparities in SUD mortality, particularly the late emergence of clusters among the black subpopulation, highlight the need for policies that address systemic biases within healthcare. National interventions should aim to improve access to culturally competent care and ensure equitable treatment for all demographic groups. This includes expanding access to medication-assisted treatment and other evidence-based therapies that have been shown to be effective in treating SUD.

Analyzing CDC mortality data is crucial for monitoring the SUD epidemic’s evolving landscape. This comprehensive data analysis reveals patterns and trends essential for understanding the epidemic’s complex dynamics. Future research should include such datasets to track changes in mortality rates, emerging SUD clusters, and demographic impact shifts. Ongoing analysis is vital for identifying new areas of concern, evaluating current intervention effectiveness, and adjusting strategies. As the SUD epidemic evolves, influenced by factors like new synthetic drugs and changing societal conditions, continuous monitoring of CDC mortality data will be invaluable for public health officials, researchers, and policymakers. This vig-ilance will enable a responsive, data-driven approach, guiding the development of targeted prevention and treatment strategies adaptable to the epidemic’s complex dynamics. However, national policies should focus on standardizing substance classification across jurisdictions to ensure accurate and consistent data collection. This would enhance the quality of mortality data and enable more precise identification of spatiotemporal clusters. Furthermore, national strategies should promote the adoption of evidence-based practices in pain management and opioid prescribing to reduce disparities and prevent misuse.

## 5 Limitations of the study

This study has two primary limitations. First, relying on mortality data classified under specific ICD-10 codes for unintentional drug poisoning as a proxy for estimating SUD mortality introduces potential inaccuracies. Cases where the substance implicated in death is not accurately identified by the medical examiner may be missed in our dataset. This underscores the need for improved quality in characterizing SUD-related deaths and standardizing substance classification across jurisdictions, including explicit coding for emerging substances like fentanyl. Second, the ecological and longitudinal nature of our analysis means that while our findings indicate broader trends, they may not uniformly apply across all scales due to the multifaceted factors influencing the SUD epidemic. Our exploratory approach, using spatial scan statistics to elucidate the spatiotemporal dynamics of SUD mortality, highlights general patterns but also necessitates further investigation into substance-specific clusters. A more detailed analysis, segmented by substance and focused on specific geographical and temporal segments, is crucial for understanding how specific substances contribute to the emergence and distribution of SUD mortality clusters. This comprehensive perspective is essential for tailoring interventions and policy responses to the complex landscape of the SUD epidemic.

## 6 Conclusions

This study has explored the spatiotemporal dynamics of the SUD epidemic in the US, revealing critical insights into the shifting epicenter of the epidemic, the populations most at risk, and the regional heterogeneity of the crisis. Our findings underscore the need for targeted public health interventions that are both timely and geographically specific. Our analysis identified 27 significant spatiotemporal clusters of elevated SUD mortality across the general population, emerging between 2005 and 2018 and persisting until 2020. These clusters highlight a dynamic shift in the spatial epicenter of the epidemic from the western to the eastern US around 2016. This geographic shift has profound public health implications, necessitating adaptive strategies that address the evolving nature of the epidemic.

The transition of the epicenter of the epidemic from the West to the East underscores the need for region-specific interventions. Early clusters in the West, particularly in rural areas, require sustained focus on reducing overdose deaths through enhanced access to treatment and harm reduction services like increasing the availability of Naloxone, and enhancing substance use education. As the epidemic has moved East, particularly into urban areas, interventions must be adapted to address the unique challenges of these densely populated regions, such as increased availability of synthetic opioids and higher population densities, that need specific responses like ensuring sufficient treatment facilities, effective needle exchange programs, and robust surveillance systems in these areas.

Our study highlights the emergence of the black population as a recent vulnerable group significantly affected by the SUD epidemic. From 2013 onwards, clusters of high mortality rates among the black subpopulation have emerged, with a notable spike in mortality from 2018 onwards. This shift indicates that public health strategies must now prioritize addressing systemic healthcare biases, ensuring equitable access to treatment, and tailoring interventions to the specific needs of black communities. This includes expanding access to medication-assisted treatment (MAT) and other evidence-based therapies. Likewise, addressing socioeconomic determinants of health and improving healthcare access in these communities are crucial steps in mitigating this emerging crisis.

The heterogeneity in the impact of the epidemic across different regions and populations underscores the importance of localized, data-driven public health interventions. The dynamic nature of the epidemic, with shifting epicenters and emerging vulnerable populations, calls for flexible and adaptive strategies. Implementing advanced surveillance systems such as FROST, SNOW, DOSE, SUDOR, and DAWN can provide timely alerts about specific substance threats, facilitating rapid and precise public health responses. Continuous monitoring of CDC mortality data and integrating broader datasets will enable the identification of new risk areas and the evaluation of current effectiveness of the intervention strategies. This will generate a responsive, data-driven approach to curve the SUD epidemic and ensure that no one is left behind.

## Data Availability

All data produced are available online at https://www.cdc.gov/nchs/nvss/nvss-restricted-data.htm

